# Detection of SARS-CoV-2 within the healthcare environment: a multicentre study conducted during the first wave of the COVID-19 outbreak in England

**DOI:** 10.1101/2020.09.24.20191411

**Authors:** Ginny Moore, Helen Rickard, David Stevenson, Paz Aranega Bou, James Pitman, Ant Crook, Katherine Davies, Antony Spencer, Chris Burton, Linda Easterbrook, Hannah E. Love, Sian Summers, Stephen R. Welch, Nadina Wand, Katy-Anne Thompson, Thomas Pottage, Kevin S. Richards, Jake Dunning, Allan Bennett

## Abstract

Understanding how Severe Acute Respiratory Syndrome Coronavirus 2 (SARS-CoV-2) is spread within the hospital setting is essential if staff are to be adequately protected, effective infection control measures are to be implemented and nosocomial transmission is to be prevented.

The presence of SARS-CoV-2 in the air and on environmental surfaces around hospitalised patients, with and without respiratory symptoms, was investigated. Environmental sampling was carried out within eight hospitals in England during the first wave of the COVID-19 outbreak. Samples were analysed using reverse transcription polymerase chain reaction (RT-PCR) and virus isolation assays.

SARS-CoV-2 RNA was detected on 30 (8.9%) of 336 environmental surfaces. Ct values ranged from 28·8 to 39·1 equating to 2·2 × 10^5^ to 59 genomic copies/swab. Concomitant bacterial counts were low, suggesting the cleaning performed by nursing and domestic staff across all eight hospitals was effective. SARS-CoV-2 RNA was detected in four of 55 air samples taken <1 m from four different patients. In all cases, the concentration of viral RNA was low and ranged from <10 to 460 genomic copies per m^3^ of air. Infectious virus was not recovered from any of the PCR positive samples analysed.

Effective cleaning can reduce the risk of fomite (contact) transmission but some surface types may facilitate the survival, persistence and/or dispersal of SARS-CoV-2. The presence of low or undetectable concentrations of viral RNA in the air supports current guidance on the use of specific PPE ensembles for aerosol and non-aerosol generating procedures.

## Introduction

Over the course of 2020, Severe Acute Respiratory Syndrome coronavirus 2 (SARS-CoV-2), the causative agent of Coronavirus disease-19 (COVID-19) has spread rapidly across the globe and, as of 15 August 2020, had infected 21 million people and caused over 750,000 deaths.^1^ Transmission of respiratory viruses can occur through inhalation of respiratory droplets (particles >5µm in diameter) and infectious aerosols (< 5µm in diameter) and/or contact with respiratory droplets either directly or indirectly via contaminated surfaces. The rapid spread of COVID-19 has led many to conclude that airborne transmission must be involved.^2^ This, though, is widely debated and, according to current evidence, SARS-CoV-2 is primarily transmitted via droplet and contact routes, although it is acknowledged, that airborne transmission could occur in specific circumstances and settings.^3^

Healthcare workers (HCWs) and others on the front line are at increased risk of acquiring infection.^4^ Medical aerosol generating procedures (AGPs), such as intubation, non-invasive ventilation and airway suctioning can produce droplets < 5µm in diameter and have been associated with increased transmission of SARS-CoV-1 from patients to HCWs.^5^ It is argued, however, that there is limited evidence to link AGPs with transmission of respiratory infections, including COVID-19.^6^ Air samples taken during tracheostomy procedures, high flow nasal oxygen treatment, non-invasive ventilation and nebulisation have not contained SARS-CoV-2 RNA^7^ and HCWs exposed to unrecognised COVID-19 patients undergoing similar high-risk AGPs have not become infected.^8^ Nonetheless, occupational exposure has resulted in infection^9^ and it has been estimated that in England, patient to HCW transmissions could be responsible for 57% of HCW infections.^10^ Nosocomial transmission may also account for 20% of infections in inpatients^10^ and so understanding how SARS-CoV-2 is spread within the hospital setting is essential to ensure staff are adequately protected and effective infection control measures are implemented.

Several studies, utilising a range of air and surface sampling methods, have been carried out to determine the presence and prevalence of SARS-CoV-2 in the healthcare environment.^11–21^ The detection of viral RNA in air samples differs with study with some reporting widespread airborne contamination^14, 18, 21^ but many reporting low or non-detectable concentrations^13, 15, 16, 19^ even in samples collected 10 cm from the face of positive patients.^12^

Surfaces frequently touched by staff and/or patients are often contaminated with bacterial pathogens. Likewise, SARS-CoV-2 RNA has been detected on high-contact surfaces such as computers, bed rails and door handles. Again, the extent of this surface contamination differs with study. Reported positivity rates range from 0·8% to over 70% with those studies reporting a comparatively higher level of airborne contamination also detecting widespread surface contamination.^18, 21^ In many cases, sampling was performed before routine cleaning but the efficacy of cleaning was not assessed.^11, 13, 15^ When comparative samples were taken, SARS-CoV-2 RNA was detected on 61% of surfaces sampled prior to cleaning but was not detected on any surface after it had been cleaned.^17^ The proportion of surfaces contaminated with viral RNA can also differ with ward type. Some studies have detected little to no surface contamination in intensive care units (ICUs) but have detected widespread contamination within general wards.^11, 21^ By contrast, others report comparatively higher positivity rates within the ICU setting.^14, 20^

Environmental sampling can provide important information about the spread of healthcare-associated infections. It is though, resource-intensive and time-consuming and, thus, many studies investigating SARS-CoV-2 and its contamination of the healthcare environment have focused on a single hospital and, in the context of the COVID-19 pandemic, a single point in time. Sampling frequency is also, in general, low, meaning that results often represent a snapshot in time and place.

In a rapid evolving outbreak, there is a need to gain quick understanding of certain trends and whilst snapshot samples by themselves cannot be considered representative, they can, when taken together, provide useful data relating to type, level and location of environmental contamination. To date, however, differences in study setting, protocol and methodology have led to inconsistency in the results obtained making it difficult to draw any firm conclusions relating to SARS-CoV-2 and its presence within the healthcare environment.

As part of the Public Health England (PHE) national incident response, the presence of SARS-CoV-2 in the air and on environmental surfaces around hospitalised patients, with and without respiratory symptoms, was investigated. Environmental sampling, utilising standard methods, was carried out within 8 acute hospital trusts in England. Trends, in terms of type and level of surface contamination and the potential for AGPs to disperse SARS-CoV-2 have been identified and these provide evidence to support current infection prevention and control guidance including the use of personal protective equipment.

## Methods

Between 03/03/20 and 12/05/20, the study team visited eight hospitals (three on more than one occasion; Figure 1) and carried out environmental sampling in areas where patients infected with SARS-CoV-2 were receiving care. These included 11 negative pressure isolation rooms, 11 neutral pressure side rooms, six ICU/HDU open cohorts and 12 non-ICU cohort bays. Whilst sampling primarily focused on 44 individual bed spaces (Table 1), samples were also taken from the wider ward environment (e.g. nursing stations; patient toilet areas) and from non-COVID wards. Medical procedures being performed and obvious symptoms such as coughing were observed and recorded. Patient details (hospital number; date of admission; date of diagnosis) were collected for future correlation with clinical virology results. Details regarding routine and terminal (discharge) cleaning were also collected.

**Figure 1:**
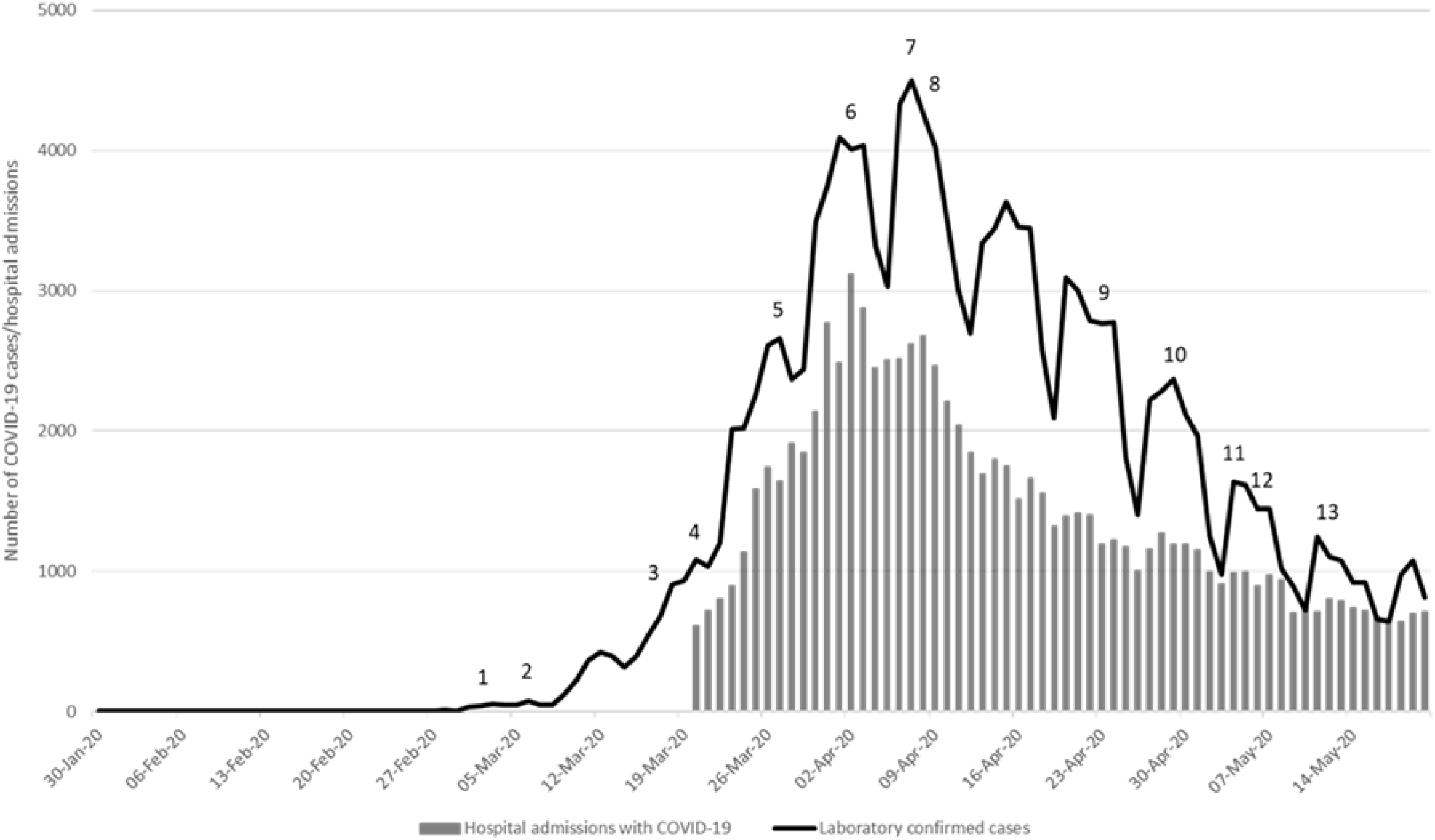
Sampling date in relation to the number of laboratory confirmed cases and hospital admission with COVID-19

**Table 1:** Sampling primarily focused on individual bed spaces (44 different patients)

| visit*<br>(hospital) | patient | ward | location | days since<br>symptom<br>onset | days since<br>admission | days since first<br>SARS-CoV-2 +<br>swab | notable<br>treatment | no. of<br>surfaces<br>'positive' for<br>SARS-CoV-2<br>RNA | SARS-CoV-2<br>RNA detected<br>in the air? |
| --- | --- | --- | --- | --- | --- | --- | --- | --- | --- |
| 1 (A) | 1 | ID | single room (-ve pressure) | 7 | 5 | 5 | none | 0 | No |
|  | 2 | ID | single room (-ve pressure) | .. | 4 | 5 | none | 0 | No |
|  | 3 | ID | single room (-ve pressure) | .. | 3 | .. | none | 0 | No |
|  | 4 | ICU | single room (-ve pressure) | 10 | 4 | 5 | O<sub>2</sub> (Venturi) | 0 | No |
| 2 (B) | 5 | ID | single room (-ve pressure) | 11 | .. | 6 | none | 0 | No |
|  | 6 | ID | single room (-ve pressure) | 13 | .. | 10 | none | 0 | No |
|  | 7 | ID | single room (-ve pressure) | 7 | .. | 3 | none | 4 | No |
| 3 (C) | 8 | ID | single room (-ve pressure) | 10 | 6 | 7 | none | 0 | No |
|  | 9 | ID | single room (-ve pressure) | 5 | 3 | 3 | O<sub>2</sub> (Venturi) | 0 | No |
|  | 10 | ID | single room (-ve pressure) | 8 | 1 | 1 | none | 0 | No |
|  | 11 | ID | single room (-ve pressure) | 10 | 3 | 3 | none | 2 | No |
| 4 (D) | 12 | ICU | cohort bay | 4 | 4 | 4 | ECMO | 0 | No |
|  | 13 | ICU | cohort bay | 9 | 9 | 1 | ECMO | 0 | No |
|  | 14 | ICU | cohort bay | 11 | 10 | 11 | ECMO | 0 | No |
|  | 15 | ICU | cohort bay | 6 | 6 | 6 | ECMO | 0 | No |
|  | 16 | ICU | cohort bay | 12 | 12 | 11 | ECMO | 0 | No |
|  | 17 | ICU | cohort bay | 17 | 17 | 17 | ECMO | 0 | No |
| 5 (D) | 18 | GW | cohort bay | 9 | 5 | 5 | O<sub>2</sub> (Venturi) | 0 | No |
|  | 19 | GW | cohort bay | 6 | 2 | 1 | O<sub>2</sub> (Venturi) | 1 | Yes |
|  | 20 | GW | cohort bay | 8 | 6 | 6 | none | 0 | Yes |
|  | 21 | GW | cohort bay | 9 | 1 | 1 | nebuliser | 0 | No |
| 6 (E) | 22 | GW | side room (neutral pressure) | 10 | 3 | 3 | CPAP | 1 | No |
|  | 23 | GW | side room (neutral pressure) | 4 | 2 | 2 | O<sub>2</sub> (Venturi) | 0 | No |
|  | 24 | ICU | side room (neutral pressure) | 17 | 7 | 7 | intubated | 0 | No |
|  | 25 | ICU | cohort bay | 9 | 4 | 3 | intubated | 0 | No |
| 7 (F) | 26 | HDU | side room (neutral pressure) | 10 | 4 | 4 | CPAP | 1 | Yes |
|  | 27 | HDU | side room (neutral pressure) | 8 | 7 | 7 | CPAP | 0 | Yes |
| 8 (G) | 28 | HDU | cohort bay | 12 | 10 | 9 | CPAP | 2 | No |
|  | 29 | HDU | cohort bay | 26 | 16 | 16 | CPAP | 0 | No |
|  | 30 | HDU | cohort bay | 19 | 12 | 12 | O<sub>2</sub> (Venturi) | 0 | No |
|  | 31 | HDU | cohort bay | 15 | 8 | 8 | CPAP | 2 | No |
| 9 (H) | 32 | ICU | side room (neutral pressure) | 9 | 4 | 4 | NIV | 0 | No |
|  | 33 | ICU | cohort bay | 7 | 2 | 2 | CPAP | 0 | No |
|  | 34 | GW | side room (neutral pressure) | .. | .. | 6 | NIV | 2 | No |
|  | 35 | ICU | cohort bay | 26 | 16 | 2 | nebuliser | 1 | No |
| 10 (H) | 36 | ICU | cohort bay | 10 | 6 | 1 | NIV | 0 | No |
|  | 37 | ICU | cohort bay | 6 | 16 | 1 | nebuliser | 0 | No |
| 11 (F) | 38 | HDU | side room (neutral pressure) | .. | .. | 2 | CPAP | 0 | No |
|  | 39 | HDU | side room (neutral pressure) | .. | .. | 4 | CPAP | 0 | No |
| 12 (H) | 40 | ICU | side room (neutral pressure) | .. | .. | .. | CPAP | 0 | No |
| 13 (D) | 41 | GW | cohort bay | .. | 34 | 34 | none | 0 | No |
|  | 42 | GW | side room (neutral pressure) | 3 | 3 | 3 | none | 3 | No |
|  | 43 | GW | cohort bay | 45 | 45 | 44 | none | 0 | No |
|  | 44 | GW | cohort bay | 14 | 14 | 13 | none | 0 | No |

\* see Figure 1
ID: infectious diseases
ICU: intensive care unit
HDU: high dependency unit
GW: general ward
CPAP: continuous positive airway pressure
NIV: non-invasive ventilation
O<sub>2</sub> (Venturi): oxygen via a Venturi mask
.. data not available/collected

Surfaces deemed to be high-contact sites were sampled using nylon flocked swabs (Copan, Brescia, Italy) wetted with universal transport medium and from 27/03/20, to provide an indication of general surface cleanliness, tryptone soya agar contact plates (Oxoid Ltd, Basingstoke, UK). Air samples were taken using two types of active air sampler: a Coriolis µ air sampler (bertin Instruments, Montigny-le-Bretonneux, France), operating at 300 L/min and collecting into 15 ml RNase-free phosphate buffered saline (PBS) and an MD8 air sampler (Sartorius, Göttingen, Germany), operating at 50 L/min and collecting onto a gelatine membrane filter. Both samplers were positioned close to patients (< 1 m) with and without respiratory symptoms and operated for 10 minutes. The type and duration of AGP, if any, was noted. Ambient temperature and relative humidity were monitored.

All samples were returned to PHE Porton Down. Agar contact plates were incubated at 37°C for 48h whilst the air and swab samples (for virus detection) were frozen at −80°C prior to processing. Laboratory-based validation experiments confirmed that neither the transport or storage conditions adversely affected subsequent RT-PCR analysis.

RNA was extracted from aliquots (140 μl) of each swab and Coriolis air sample using the QIAamp Viral RNA Mini Kit (Qiagen Ltd, Manchester, UK). The remaining Coriolis sample was concentrated to < 1 ml using a Vivaspin® 20 centrifugal concentrator. Each gelatine membrane was dissolved in 10 ml Minimum Essential Medium (MEM). Aliquots (140 μl) of both were extracted.

In total 425 samples (surface swabs (n=336) and air (n=89)) were analysed for SARS-CoV-2 using RT-PCR. All samples were screened in duplicate using one of the following targets: RNA dependent RNA polymerase (RdRp) with probe 2, Envelope (E) or Nucleocapsid (N) and ORF1ab (Viasure, CerTest Biotec, Zaragoza). A sample was considered positive when amplification was detected in both replicates or ‘suspect’ when it was detected in only one. ‘Suspect’ samples were re-analysed and considered positive if amplification was detected in both replicates. All positive samples were quantified using the N target on the Viasure platform. Amplification in one replicate was considered sufficient for quantification. Samples that could not be quantified were re-extracted and quantification reattempted.

Virus isolation was carried out on all positive samples with a Ct value < 34. Vero E6 cells (Vero C1008; ATCC CRL-1586) in culture medium (MEM supplemented with GlutaMAX™-I, 10% (v/v) fetal bovine serum (FBS), 1X (v/v) non-essential amino acids and 25 mM HEPES) were incubated at 37°C. Cells (1 × 10^6^ cells/25 cm^2^ flask) were washed with 1X PBS and inoculated with ≤1 ml environmental sample and incubated at 37°C for 1 hour. Cells were washed with 1X PBS and maintained in 5 ml culture medium (4% FBS) with added antibiotic-antimycotic (4X), incubated at 37°C for 7 days and monitored for cytopathic effects (CPE). Cell monolayers that did not display CPE were sub-cultured a maximum of three times, providing continuous cultures of ~30 days.

## Results

Environmental sampling was carried out in and around the bed space of 44 different patients, 35 (80%) of whom were male (Table 1). Twenty-three patients had been admitted to intensive care (n=15) or a respiratory high dependency unit (n=8) whilst 21 patients occupied beds in a non-ICU setting. These included 10 patients who, after being diagnosed early in the outbreak, were admitted to infectious diseases units. At the time of sampling, 21 patients were receiving mechanical ventilation either invasively (n=8) or non-invasively (n=13), six patients were receiving oxygen via a Venturi mask and three patients required drugs or saline to be administered by nebulisation. All patients had tested positive for SARS-CoV-2 and the median time since diagnosis was 5 days (range 1 – 44 days). Time since symptom onset ranged from 3 to 45 days.

In total, 336 surfaces were sampled for bacteria and/or SARS-CoV-2. The mean aerobic colony count was 1 cfu/cm^2^. Of those surfaces with more extensive bacterial contamination (>2·5 cfu/cm^2^), 18 (70%) were associated with a patient’s bed (bed rail, bed control and/or nurse call button) or mobile phone. SARS-CoV-2 RNA was detected on 30 (8·9%) of the 336 surfaces sampled (Table 2). Of the 44 individual bed spaces, 10 were contaminated with viral RNA and accounted for 19 (63%) of all positive sites. In addition to nurse call buttons (n = 4), bed control panels (n = 3) and mobile phones (n = 3), viral RNA was also detected on bedside equipment (e.g. monitor screens, syringe drivers and computer keyboards) particularly in the ICU/HDU setting. However, in the non-ICU setting, 27% of surfaces contaminated with SARS-CoV-2 RNA were located outside the patient bed area. These included toilet door handles and portable vital signs monitors which together accounted for 26% of all positive sites.

**Table 2:** SARS-CoV-2 was detected on 30 of 336 surfaces sampled across eight acute hospital Trusts. All positive samples were quantified using the N target on the Viasure platform (CerTest Biotec, Zaragoza).

| Sample location |  | Surface sampled | mean Ct value | mean genomic copies/swab |
| --- | --- | --- | --- | --- |
| general ward | wider ward | toilet door handle | 28.80 | $2.19 \times 10^5$ |
| general ward | wider ward | toilet door handle | 38.94 | $1.90 \times 10^2$ |
| general ward | cohort bay | toilet door handle | 38.16 | $1.56 \times 10^2$ |
| infectious diseases | isolation room | toilet door handle | 38.45 | $3.72 \times 10^2$ |
| general ward | side room | door handle | 37.95 | $9.94 \times 10^1$ |
| general ward | side room | nurse call button | 30.71 | $2.89 \times 10^4$ |
| infectious diseases | isolation room | nurse call button | 33.30 | $9.80 \times 10^3$ |
| general ward | side room | nurse call button | 36.21 | $1.27 \times 10^3$ |
| HDU | side room | nurse call button | 36.26 | $1.26 \times 10^3$ |
| HDU | wider ward | portable vital signs monitor | 35.89 | $1.58 \times 10^3$ |
| general ward | cohort bay | portable vital signs monitor | 36.70 | $9.03 \times 10^2$ |
| general ward | cohort bay | portable vital signs monitor | 37.82 | $4.17 \times 10^2$ |
| general ward | cohort bay | portable vital signs monitor | 38.97 | $1.87 \times 10^2$ |
| infectious diseases | isolation room | mobile phone | 30.34 | $7.49 \times 10^4$ |
| general ward | cohort bay | mobile phone | 36.98 | $4.15 \times 10^2$ |
| general ward | cohort bay | mobile phone | 37.26 | $3.08 \times 10^2$ |
| general ward | side room | bed rail | 35.56 | $1.01 \times 10^3$ |
| infectious diseases | isolation room | bed control | 35.12 | $2.76 \times 10^3$ |
| general ward | cohort bay | bed control | 38.10 | $3.43 \times 10^2$ |
| HDU | cohort bay | bed control | 38.92 | unable to quantify* |
| HDU | cohort bay | monitor | 35.72 | $8.97 \times 10^2$ |
| HDU | cohort bay | monitor | 36.11 | $7.41 \times 10^2$ |
| HDU | cohort bay | syringe driver | 37.02 | $3.64 \times 10^2$ |
| ICU | cohort | bedside computer | 39.11 | $5.91 \times 10^1$ |
| general ward | side room | bedside computer | 38.71 | unable to quantify* |
| infectious diseases | isolation room | chair arm | 37.84 | $4.23 \times 10^2$ |
| general ward | cohort bay | curtain | 37.98 | $3.72 \times 10^2$ |
| general ward | side room | window sill | 38.05 | $7.63 \times 10^1$ |
| infectious diseases | isolation room | air vent | 37.52 | $2.75 \times 10^2$ |
| A&E | Resus bay | trolley drawer | 37.89 | $8.66 \times 10^1$ |

\* SARS-CoV-2 detected on initial screening but quantification was unsuccessful
ICU: intensive care unit
HDU: high dependency unit
A&E: accident and emergency

RT-PCR Ct (cycle threshold) values ranged from 28·8 to 39·1 which when quantified equated to 2·2 × 10^5^ to 59 genomic copies per swab. Samples with a Ct value < 34 were incubated on Vero E6 cells. No CPE or a decrease in Ct values across the course of three serial passages were observed suggesting the samples did not contain infectious virus.

Ambient temperature and relative humidity differed with ward and ranged from 21°C to 25°C and from 21% to 41% respectively. Air samples were collected using two types of high-volume air sampler but SARS-CoV-2 RNA was only detected in four (7·3%) of the 55 samples taken using the Coriolis μ sampler. Two of these samples were taken in two different single rooms (neutral pressure). In both cases, the sampler was positioned close (< 1 m) to a patient being treated with Continuous Positive Airway Pressure (CPAP) via a mask that covered the nose and mouth. Time since diagnosis was 4 and 7 days with both patients reporting symptoms at least 8 days prior to sampling.

Viral RNA was also detected in two air samples taken in two 4-bed cohort bays. On one of these occasions, the air sampler was positioned close to a patient who was receiving oxygen via a Venturi mask. This patient had tested positive for SARS-CoV-2 the previous day with a Ct value of 21·35. The second patient, diagnosed 6 days earlier (Ct value of 17·68), was not receiving any notable treatment. However, approximately 30-40 minutes before sampling was carried out, there was a ‘crash call’ elsewhere within the bay. There was no intubation or CPR but a significant increase in staff activity was observed and may have facilitated dispersal of airborne particles.

The total volume of each air sample was 3 m^3^ and the associated Ct values ranged from 37 to 39 which, when quantified, equated to 460 to < 10 genomic copies per m^3^ of air.

## Discussion

When sampling the healthcare environment, many variables can impact the results obtained. This can make interpretation of the data difficult, particularly if a frame of reference is lacking. In this study and to provide context, agar contact plates were used to provide an aerobic bacterial colony count and an indication of surface cleanliness. Whilst no microbiological standards exist for healthcare surfaces, a benchmark of < 2·5 cfu/cm^2^ has been suggested.^22^ SARS-CoV-2 was detected on 30 (8·9%) of the 336 surfaces sampled (Table 2). The proportion of surfaces positive for viral RNA differed with hospital and ranged from 0-27%. This likely reflects the fact that sampling was carried out on different types of ward occupied by different types of patient requiring different types of care and/or treatment (Table 1). Overall, however, the results are similar to those of other studies^13, 15, 20^ and suggest that, whilst SARS-CoV-2 can contaminate healthcare surfaces, widespread contamination is unlikely.^17^ The bacterial load on the majority (89%) of surfaces sampled was < 2·5 cfu/cm^2^ suggesting that in general and despite increased pressure on beds and workload, the routine cleaning performed by the nursing and domestic staff across all eight hospitals was effective.

Nonetheless, contamination of the healthcare environment can occur and SARS-CoV-2 RNA was detected on the same type of surface across multiple hospitals (Table 2) implying that, despite the effectiveness of the cleaning protocols employed, some types of surface could facilitate the survival, persistence and/or dispersal of SARS-CoV-2.

Patients consider the nurse call button a direct conduit to care and many patients were observed to hold the button close even whilst dozing. Intensity and frequency of contact can increase microbial transfer from hands to surface^23^ and SARS-CoV-2 RNA was detected on 4 (17%) of the nurse call buttons sampled. Ct values ranged from 30·8 to 36·2 equating to 2·9 × 10^4^ to 1·2 × 10^3^ genomic copies/swab (Table 2).

To reduce the risk of transmission of SARS-CoV-2 in the hospital setting, it is recommended that surfaces such as over-bed tables, bed rails and nurse call buttons are cleaned at least twice daily.^24^ The median number of bacteria recovered from nurse call buttons was 50 cfu/25cm^2^. Comparatively fewer bacteria (< 1 cfu/cm^2^) were recovered from tables and bed rails suggesting these surfaces are (and can be) effectively cleaned. Heavy contamination of the nurse call button has been described previously^25^ and staff should be reminded that routine cleaning should include all aspects of the patient bed. Future consideration should be given to design modification and/or improving the ability to clean nurse call buttons.

Patient mobility can contribute greatly to the spread of bacteria within a ward.^25^ Similarly, SARS-CoV-2 RNA has been detected on patient-contact sites outside the immediate bed space ^13, 17^ and, in the current study, outside of cohort bays; specifically, toilet door handles. The presence of SARS-CoV-2 RNA on door handles has been reported previously ^13, 14, 20^ and the contact area between the hand and handle and the pressure of grip likely facilitates transfer to and from the hands. In this study, the amount of SARS-CoV-2 RNA detected on one door handle was 2·2 × 10^5^ genomic copies/swab implying significant transfer from a contaminated hand. Despite this, we were unable to culture viable virus. The lowest genomic copy number (N gene) required to isolate virus from clinical samples is reportedly 5 × 10^5^ genomic copies/ml^26^; higher than the copy number in any of the environmental samples collected during this study. Subjecting the samples to multiple freeze-thaw cycles may also have impacted infectivity by disrupting virion and genome integrity^26^. Regardless, there is potential for viable virus to contaminate a single door handle and to be transferred to the hands of numerous successive contacts and as a consequence, to other inanimate surfaces.^27^

SARS-CoV-2 RNA was detected on 4·9% (7/143) and 13·8% (22/159) of the surfaces sampled in the ICU/HDU and non-ICU wards respectively. In contrast to patients admitted to cohort wards, patients in ICUs/HDUs are more likely to be bed bound and be receiving mechanical ventilation. Reduced patient mobility likely contributed to the less frequent detection of SARS-CoV-2. However, viral RNA was still detected on staff contact sites (e.g. monitor screens, syringe drivers; Table 2).

Disposable gloves are an important element of PPE and can prevent the hands of HCWs from acquiring pathogens. However, during routine patient care, the glove surface itself can become contaminated. If gloves are not regularly and appropriately changed then contamination of surfaces via gloved hands can occur.^19^ When caring for COVID-19 patients, particularly in the ICU/HDU setting, the requirement to don full PPE presents additional challenges in terms of preserving PPE and ensuring staff know how to implement appropriate hand hygiene within an outbreak setting.^14, 19^

Non-critical medical devices (e.g. blood pressure cuffs and temperature probes) have been implicated in nosocomial infection.^28^ SARS-CoV-2 was detected on four (31%) of 13 portable vital signs monitors (Table 2). The highest level of viral RNA (1·6 × 10^3^ genomic copies/swab) was detected on a fingertip pulse-oximeter associated with a machine that had been removed from a single room occupied by a COVID-positive patient. The other three machines were located on cohort bays. When (or on whom) these machines were last used or when they were last cleaned is not known and results demonstrate the presence and/or persistence of viral RNA and not infectious virus. Nonetheless, contact pressure has been shown to significantly affect viral transfer to and from fingerpads^29^ and in the absence of cleaning, fingertip pulse-oximeters could facilitate transmission of SARS-CoV-2, particularly between asymptomatic and non-infected patients.

SARS-CoV-2 RNA was detected in four (7·3%) of the high volume (3 m^3^) air samples taken using the Coriolis sampler. It is not known what may have contributed to this airborne contamination but two of these samples were taken < 1 m from two patients receiving CPAP therapy (Table 1). CPAP is considered an AGP. However, air samples were taken close to 11 other patients receiving non-invasive ventilation (NIV), 7 of whom had also tested positive < 7 days earlier. No viral RNA was detected. The make/model of CPAP machine used to treat these two patients was not used elsewhere and it is possible that the equipment used to deliver NIV to patients may promote the generation and/or release of aerosols.^30^ How the apparatus is used or tolerated may also have an effect. During sampling, one of the two patients was observed to turn in bed - multiple times and on one occasion disconnect the CPAP machine to aid movement.

The dispersal distance of exhaled air from a jet nebulizer and Venturi-type oxygen mask is estimated to be 0.8 m and 0.4 m respectively.^30^ In this study, SARS-CoV-2 was not detected in any air sample collected during drug nebulisation. Viral RNA was detected < 1 m from one (of six) patients receiving oxygen. Time since diagnosis and symptom onset was 1 and 6 days respectively; comparatively earlier than many of the other patients (Table 1). Others hypothesise that the concentration of SARS-CoV-2 in the air and/or on high-touch surfaces is highest during the first week of illness,^11^ suggesting that new admissions to hospital may have greater potential to transmit the virus to others. It has been suggested that placing suspected COVID-19 patients in single rooms or bays that are fully disinfected between admissions could reduce nosocomial infection rates by 80%.^10^

In all four cases where SARS-CoV-2 was detected in air samples, the concentration of viral RNA was low and ranged from 460 to < 10 genomic copies per m^3^ of air. As discussed, samples containing this level of viral nucleic acid are unlikely to contain viable (infectious) virus^27^ and this finding, together with the inability to detect SARS-CoV-2 RNA in all other air samples, supports current guidance on the use of specific PPE ensembles for aerosol- and non-aerosol generating procedures. It is acknowledged, however, that many of the procedures believed to generate aerosols and droplets were not captured during this study and that samples were only collected over a 10-minute period. Unprotected, prolonged exposure to an infected patient has been linked to transmission.^9^

In a rapidly evolving outbreak situation, there is a need to gain quick understanding of certain trends; in this case, the contamination of the healthcare environment. Despite its limitations, this multi-centre study supports the findings of others^13, 15, 19-20^ and should provide assurance to HCWs. SARS-CoV-2 may be present on frequently touched surfaces but effective cleaning should reduce the risk of fomite-transmission^21^ and limit the concentration of SARS-CoV-2 in aerosols.^17^ Recommendations to regularly clean frequently touched surfaces are warranted and the need to clean items such as door handles, nurse call buttons and multi-use patient monitoring equipment should be emphasised. In wards caring for COVID-19 patients, viral RNA in the air was either not detected or was present only at a very low concentration. Our results suggest that if worn and used correctly, the PPE recommended in the UK, including components to protect against aerosol exposures when indicated, should provide adequate protection against the potential virus exposure risks identified in this study.

## Data Availability

The corresponding author collected and has access to all study data

## Acknowledgements

We thank the staff and patients at the hospitals that participated in this study. The views expressed in this article are those of the author(s) and are not necessarily those of Public Health England or the Department of Health and Social Care.

## Contributors

GM and HR carried out the environmental sampling, collected, analysed and interpreted the data and drafted the manuscript. AC, AS, KAT and TP formed part of the sampling team. JP, KD, CB, LE, SS, SW, HL, KR, DS and PAB were responsible for sample processing, inactivation and nucleic acid extraction. HL, KR, DS, PAB and NW were responsible for PCR and analysis. KR, SS and CB were responsible for virus isolations. KAT carried out validation studies. TP was responsible for logistics. JD and AB contributed to study conception and design. JD provided clinical input. All authors read and approved the final manuscript.

